# TMPRSS2 Expression in Lung Tissue of Prostatic Adenocarcinoma Patients: A Pathologic Perspective on Androgen Deprivation Therapy

**DOI:** 10.1101/2025.05.03.25326931

**Authors:** Marcela Riveros Angel, David Loeffler, Ahmad Charifa, Ryan B. Sinit, Taylor Amery, Beyza Cengiz, Tomasz M. Beer, George V. Thomas

**Author notes:** The authors have no relevant financial interest in the products or companies described in this article. Corresponding author: George V. Thomas, MD, Department of Pathology & Laboratory Medicine, Oregon Health & Science University, 3181 SW Sam Jackson Park Rd, Portland, OR 97239.

## Abstract

**Context:** Severe acute respiratory syndrome coronavirus 2 (SARS-CoV-2) cellular entry is facilitated by transmembrane protease serine 2 (TMPRSS2), which is regulated by the androgen receptor (AR). Androgen deprivation therapy (ADT), widely used in prostate cancer treatment, may potentially modulate TMPRSS2 expression, affecting SARS-CoV-2 infection susceptibility and severity.

**Objective:** To evaluate the impact of ADT on pulmonary TMPRSS2 expression in prostate cancer patients and analyze differences in expression patterns associated with specific ADT regimens.

**Design:** We examined TMPRSS2 immunohistochemical expression in lung tissue from 20 consecutive autopsy cases of men with prostate cancer (6 receiving ADT at time of death), compared with non-ADT prostate cancer patients and age-matched women controls. Histoscores were calculated by assessing percentage and intensity of pneumocyte TMPRSS2 expression.

**Results:** Prostate cancer patients receiving ADT showed significantly reduced pulmonary TMPRSS2 expression compared to non-ADT patients (mean histoscores: 152.7 vs. 225.0, p=0.037) and age-matched women controls (mean histoscores: 152.7 vs. 238.0, p=0.024). Direct AR antagonists (apalutamide, bicalutamide) produced more pronounced TMPRSS2 suppression than GnRH modulators or androgen biosynthesis inhibitors. No significant correlation was observed between TMPRSS2 expression and Gleason score, PSA levels, or underlying lung pathology.

**Conclusion:** Our findings demonstrate that ADT significantly reduces pulmonary TMPRSS2 expression, with direct AR antagonists showing the strongest effect. This suggests a potential mechanistic explanation for differential COVID-19 susceptibility and provides rationale for investigating AR-targeted therapies as potential protective interventions against SARS-CoV-2 infection severity.

## Introduction

The coronavirus disease 2019 (COVID-19) pandemic, caused by severe acute respiratory syndrome coronavirus 2 (SARS-CoV-2), continues to pose significant global health challenges despite advances in prevention and treatment. According to the World Health Organization, COVID-19 has resulted in over 7 million deaths worldwide as of early 2025, highlighting the ongoing importance of understanding viral pathogenesis and identifying effective interventions.

SARS-CoV-2 cellular entry involves a coordinated multi-step process. The viral spike (S) glycoprotein undergoes initial priming by furin, followed by binding to the angiotensin-converting enzyme 2 (ACE2) receptor on host cell surfaces.^1,2^ Upon ACE2 binding, two distinct entry pathways are utilized: the primary pathway involves cleavage of the S2’ subunit by transmembrane protease serine 2 (TMPRSS2), exposing the viral fusion peptide and facilitating membrane fusion; alternatively, in cells with low TMPRSS2 expression, the virus can be internalized via endocytosis, with cathepsin L mediating S protein cleavage within endosomes.^1,3–6^ This mechanistic understanding has prompted investigations into targeting these entry factors as potential therapeutic approaches.

TMPRSS2, a key facilitator of SARS-CoV-2 entry, is transcriptionally regulated by the androgen receptor (AR) and is expressed in multiple tissues including prostate, lung, colon, and kidney.^7^ This androgen dependence has attracted particular attention given the significant sex disparity in COVID-19 mortality, with men experiencing approximately 1.5-fold higher death rates across all age groups. In prostate cancer, TMPRSS2 gene alterations serve as important biomarkers, with TMPRSS2-ETS gene fusions (particularly TMPRSS2-ERG) occurring in approximately 50% of cases and associated with aggressive disease features.^8–15^ While the complete implications of these fusions remain incompletely understood, they contribute to tumor invasion, angiogenesis, and androgen independence.

Androgen receptor signaling drives prostate cancer progression, making androgen deprivation therapy (ADT) the foundation of treatment for metastatic disease. Contemporary ADT approaches include GnRH analogs (leuprolide, degarelix), direct AR antagonists (bicalutamide, enzalutamide, apalutamide, darolutamide), and androgen biosynthesis inhibitors (abiraterone).^16–18^ The connection between AR-regulated TMPRSS2 expression and SARS-CoV-2 entry has prompted investigations into whether ADT might confer protection against COVID-19 severity in prostate cancer patients, though results have been inconsistent.^19^

Recent work by Schuler et al. demonstrated developmental increases in pulmonary TMPRSS2 expression, with prominent expression in secretory, ciliated, and alveolar type 1 epithelial cells. Their analysis of COVID-19 autopsy specimens revealed higher viral infection rates in TMPRSS2-expressing cells, further supporting its role in viral pathogenesis.^4^ Similarly, Samuel et al. identified associations between elevated free androgen levels and COVID-19 complications in autopsy studies.^5^

The present study was designed to determine whether ADT modulates TMPRSS2 expression in lung tissue of prostate cancer patients and to assess whether specific ADT regimens differ in their effects on pulmonary TMPRSS2 levels. We hypothesized that ADT would reduce TMPRSS2 expression in lung tissue, potentially providing a mechanistic explanation for differential COVID-19 susceptibility. Through immunohistochemical analysis of autopsy specimens, we demonstrate significant reduction in pulmonary TMPRSS2 expression in ADT-treated patients compared to untreated prostate cancer patients and controls, with direct AR antagonists showing particularly potent suppressive effects. These findings offer novel insights into the regulation of this key viral entry factor and suggest potential therapeutic strategies for mitigating SARS-CoV-2 infection severity.

## Materials and Methods

### Patient Selection and Data Collection

Following institutional review board approval, we conducted a retrospective analysis of consecutive autopsy cases from our institution (Oregon Health & Science University) between 2010 and 2019. Using the Epic SlicerDicer tool, we identified 20 male patients with documented prostatic adenocarcinoma. For each case, we retrieved comprehensive clinical information including demographics, medical history, treatment details, and laboratory values from electronic health records.

We selected lung tissue slides from the pathology archives for assessment. Exclusion criteria included completely autolyzed tissue or pneumonia-involved tissue lacking normal lung parenchyma as control. We established a control group of 10 age-matched women patients who underwent autopsy during the same period, enabling comparative analyses while accounting for age-related histological changes.

Clinical parameters documented for all cases included age at death, race, smoking history, comorbidities, cause of death, and medications. For prostate cancer patients, we additionally recorded Gleason score, treatment history, PSA levels at time of death, LDH, serum testosterone, and ADT regimen, as summarized in Table 1.

**Table 1.** Patient Demographics and Clinical Characteristics.

| **Characteristic** | **ADT Group**<br>**(n=6)** | **Non-ADT Group**<br>**(n=10)** | **Uncertain ADT**<br>**Status (n=4)** | **Women**<br>**(n=10)** |
| --- | --- | --- | --- | --- |
| **Age (years)** |  |  |  |  |
| Median (range) | 78 (60-83) | 68.5 (45-86) | 64 (61-74) | 66 (46-86) |
| **Race** |  |  |  |  |
| White | 6 (100%) | 10 (100%) | 4 (100%) | 10 (100%) |
| **Smoking History** |  |  |  |  |
| Never smoker | 3 (50%) | 4 (40%) | 1 (25%) | 3 (30%) |
| Former smoker | 3 (50%) | 5 (50%) | 2 (50%) | 5 (50%) |
| Current smoker | 0 (0%) | 1 (10%) | 1 (25%) | 2 (20%) |
| **Gleason Score** |  |  |  |  |
| 6 (3+3) | 1 (16.7%) | 7 (70%) | 0 (0%) | N/A |
| 7 (3+4) | 1 (16.7%) | 0 (0%) | 0 (0%) | N/A |
| 7 (4+3) | 1 (16.7%) | 1 (10%) | 0 (0%) | N/A |
| 9 (4+5) | 1 (16.7%) | 0 (0%) | 0 (0%) | N/A |
| 9 (5+4) | 1 (16.7%) | 1 (10%) | 0 (0%) | N/A |
| Unknown | 1 (16.7%) | 1 (10%) | 4 (100%) | N/A |
| **PSA at death (ng/mL)** |  |  |  |  |
| Median (range) | 149.6 (13.8-2308.5) | N/A | N/A | N/A |
| **ADT Regimen** |  |  |  |  |
| Leuprolide +<br>Apalutamide | 1 (16.7%) | N/A | N/A | N/A |
| Leuprolide +<br>Bicalutamide | 2 (33.3%) | N/A | N/A | N/A |
| Degarelix + Orteronel | 1 (16.7%) | N/A | N/A | N/A |
| Leuprolide + Abiraterone | 1 (16.7%) | N/A | N/A | N/A |
| Leuprolide monotherapy | 1 (16.7%) | N/A | N/A | N/A |
| **Metastatic Status** |  |  |  |  |
| Bone | 6 (100%) | 0 (0%) | 0 (0%) | N/A |
| Lymph node | 4 (66.7%) | 0 (0%) | 0 (0%) | N/A |
| Other organs | 2 (33.3%) | 0 (0%) | 0 (0%) | N/A |
| **Primary Cause of**<br>**Death** |  |  |  |  |
| Metastatic disease | 2 (33.3%) | 0 (0%) | 0 (0%) | 0 (0%) |
| Hemorrhagic<br>complications | 2 (33.3%) | 2 (20%) | 0 (0%) | 1 (10%) |
| Cardiovascular event | 1 (16.7%) | 5 (50%) | 2 (50%) | 5 (50%) |
| Respiratory<br>failure/pneumonia | 0 (0%) | 1 (10%) | 0 (0%) | 3 (30%) |
| Other | 1 (16.7%) | 2 (20%) | 2 (50%) | 1 (10%) |

### Histopathologic Evaluation

Two independent board-certified surgical pathologists, blinded to clinical data, performed thorough assessment of pathological alterations in all specimens. Pathological findings were documented and cross-referenced with diagnoses from clinical records and autopsy reports. This comprehensive approach allowed correlation of microscopic features with clinical information, including metastases, primary lung pathology, and other significant findings (Table 2).

**Table 2.** Summary of Pulmonary Histopathologic Features.

| **Histopathologic Finding** | **ADT Group**<br>**(n=6)** | **Non-ADT Group**<br>**(n=14)** | **Women Controls**<br>**(n=10)** |
| --- | --- | --- | --- |
| **Emphysematous changes** | 4 (66.7%) | 7 (50%) | 4 (40%) |
| **Vascular congestion** | 4 (66.7%) | 5 (35.7%) | 3 (30%) |
| **Pneumonia** | 2 (33.3%) | 5 (35.7%) | 6 (60%) |
| **Hemorrhage** | 0 (0%) | 3 (21.4%) | 3 (30%) |
| **Pulmonary tumor** | 2 (33.3%) | 1 (7.1%) | 0 (0%) |
| **Vascular abnormalities** | 2 (33.3%) | 0 (0%) | 1 (10%) |
| **Fibrosis** | 2 (33.3%) | 1 (7.1%) | 0 (0%) |
| **Normal lung tissue** | 0 (0%) | 1 (7.1%) | 1 (10%) |

### Immunohistochemical Analysis

Immunohistochemical staining was performed on 4 μm formalin-fixed, paraffin-embedded tissue sections using anti-TMPRSS2 antibody (clone EPR3861, dilution 1:6000; ab92323, Abcam) on a Benchmark-Ultra automated system (Ventana-Roche, Tucson, AZ, USA). The specificity of the antibody was validated using appropriate positive and negative controls.

### TMPRSS2 Expression Assessment

TMPRSS2-stained slides were digitally scanned at high resolution and independently analyzed by two pathologists blinded to clinical data. For each specimen, the percentage of pneumocyte staining was assessed, and nuclear staining intensity was categorized on a scale of 0 to 3 (0=none, 1=weak, 2=moderate, 3=strong). A histoscore was calculated for each sample using the formula: (1 × [% cells with 1+ staining] + 2 × [% cells with 2+ staining] + 3 × [% cells with 3+ staining]), yielding scores ranging from 0-300, with higher values indicating greater TMPRSS2 expression.

### Statistical Analysis

Independent samples t-tests were used to compare mean histoscores between groups. P-values <0.05 were considered statistically significant. Statistical analyses were performed using SPSS version 25.0 (IBM Corp., Armonk, NY).

## Results

### Patient Characteristics

Our cohort was comprised of 20 deceased men with prostate adenocarcinoma, all identified as white, with ages ranging from 45-86 years (median 68.5). Smoking history varied across the cohort, including never-smokers, former smokers, and current smokers. All patients presented with comorbidities, most commonly hypertension, diabetes mellitus, and cardiovascular disease (Table 1).

Of the 20 prostate cancer patients, 6 were receiving ADT at time of death (leuprolide, bicalutamide, degarelix, orteronel, apalutamide, or abiraterone), ranging in age from 60-83 years (median 78). All 6 had disseminated metastatic castration-resistant prostate cancer (CRPC), with bone being the most common metastatic site (n=6), followed by lymph nodes (n=4) and other organs (n=2). Common causes of death included hemorrhagic complications (n=2), cardiovascular events (n=1), bowel obstruction (n=1), and direct complications of metastatic disease (n=2).

Among the 14 remaining patients, 10 had no documented history of ADT, while ADT status could not be confirmed for 4 patients. This non-ADT group ranged in age from 45-86 years (median 68.5) with median Gleason Score 6 (Grade Group 1). Primary causes of death included cardiovascular/cerebrovascular events, traumatic hemorrhage, and respiratory complications.

The women control group (n=10) ranged in age from 46-86 years (median 66), with cardiovascular disease being the most common cause of death.

### Pulmonary Histopathologic Findings

Pathological examination revealed that two ADT-treated patients who died from metastatic CRPC exhibited pulmonary infiltrates of prostatic adenocarcinoma, manifesting as intravascular tumor thrombi (Figure 1A). While these findings likely contributed to clinical deterioration, they were not deemed the principal cause of death. Additional pulmonary findings in the ADT cohort included emphysematous changes, vascular congestion, multifocal pneumonia, arterial thickening, and fibrin thrombus formation (Figure 1B).

**Figure 1.**
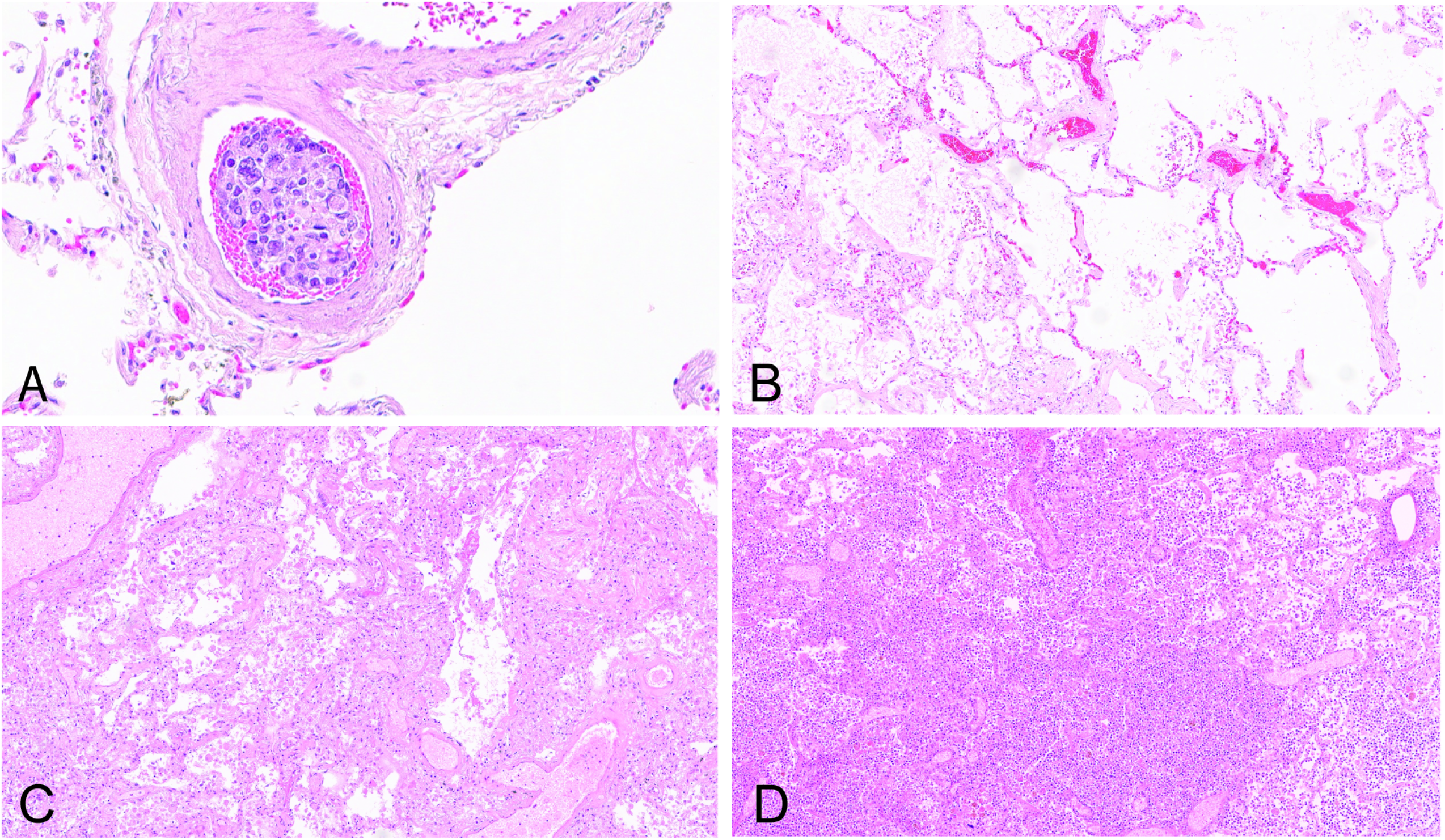
Representative pulmonary histopathology (hematoxylin-eosin stain). A, Pulmonary infiltrates of metastatic prostatic adenocarcinoma with tumor emboli in vascular spaces. B, Emphysematous changes with enlarged distal airspaces and alveolar septal destruction. C, Acute organizing pneumonia with alveolar macrophage accumulation and fibrin deposition. D, Bronchopneumonia with neutrophilic infiltration and proteinaceous exudates (original magnification: A ×200, B-D ×100).

Patients without documented ADT presented diverse pulmonary histopathology, most commonly emphysematous changes and congestion (5 out of 14), followed by inflammatory and infectious features (5 out of 14) including alveolar macrophage accumulation, acute pneumonia, and bronchial bacterial/fungal pneumonia. Bronchopneumonia was the adjudicated cause of death in one case (Figure 1C).

In the control group, 8 out of 10 subjects showed underlying pulmonary disease at time of death, with bronchopneumonia being most frequent (Figure 1D).

### TMPRSS2 Expression Analysis

Immunohistochemical analysis revealed TMPRSS2 expression primarily in alveolar pneumocytes, with varying intensity across patient groups. Quantitative assessment showed significantly reduced TMPRSS2 protein expression in prostate cancer patients receiving ADT compared to those not receiving ADT (mean histoscores: 152.7 vs. 225.0, p=0.037) (Figure 2A-D). When including patients with uncertain ADT status, the trend persisted but narrowly missed statistical significance (p=0.058).

**Figure 2.**
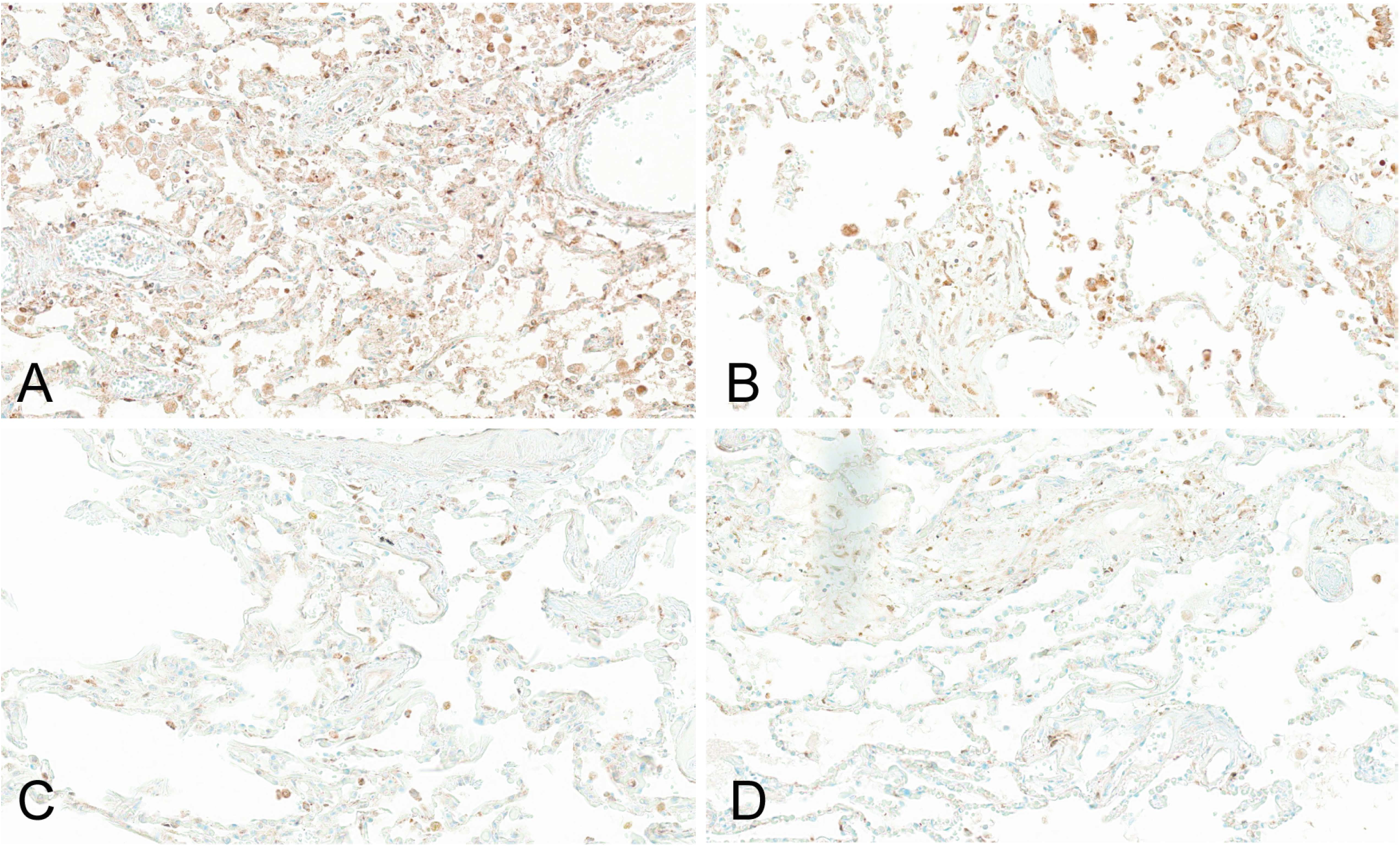
TMPRSS2 immunohistochemistry in lung tissue. A-B, Lung tissue from prostate cancer patients not treated with ADT showing strong TMPRSS2 expression (histoscores 221.1 and 277.5, respectively). C-D, Lung tissue from ADT-treated prostate cancer patients showing markedly reduced TMPRSS2 expression (histoscores 90.0 and 107.5, respectively) (original magnification ×200). **Abbreviations:** TMPRSS2, transmembrane protease serine 2.

Notably, TMPRSS2 expression was also significantly lower in ADT-treated patients compared to women controls (mean histoscores: 152.7 vs. 238.0, p=0.024), while expression was comparable between non-ADT men and women controls (p=0.164) (Figure 3). Patients with uncertain ADT status also displayed relatively low TMPRSS2 expression (mean: 136.9), potentially indicating a mixed population including some previously treated individuals.

**Figure 3.**
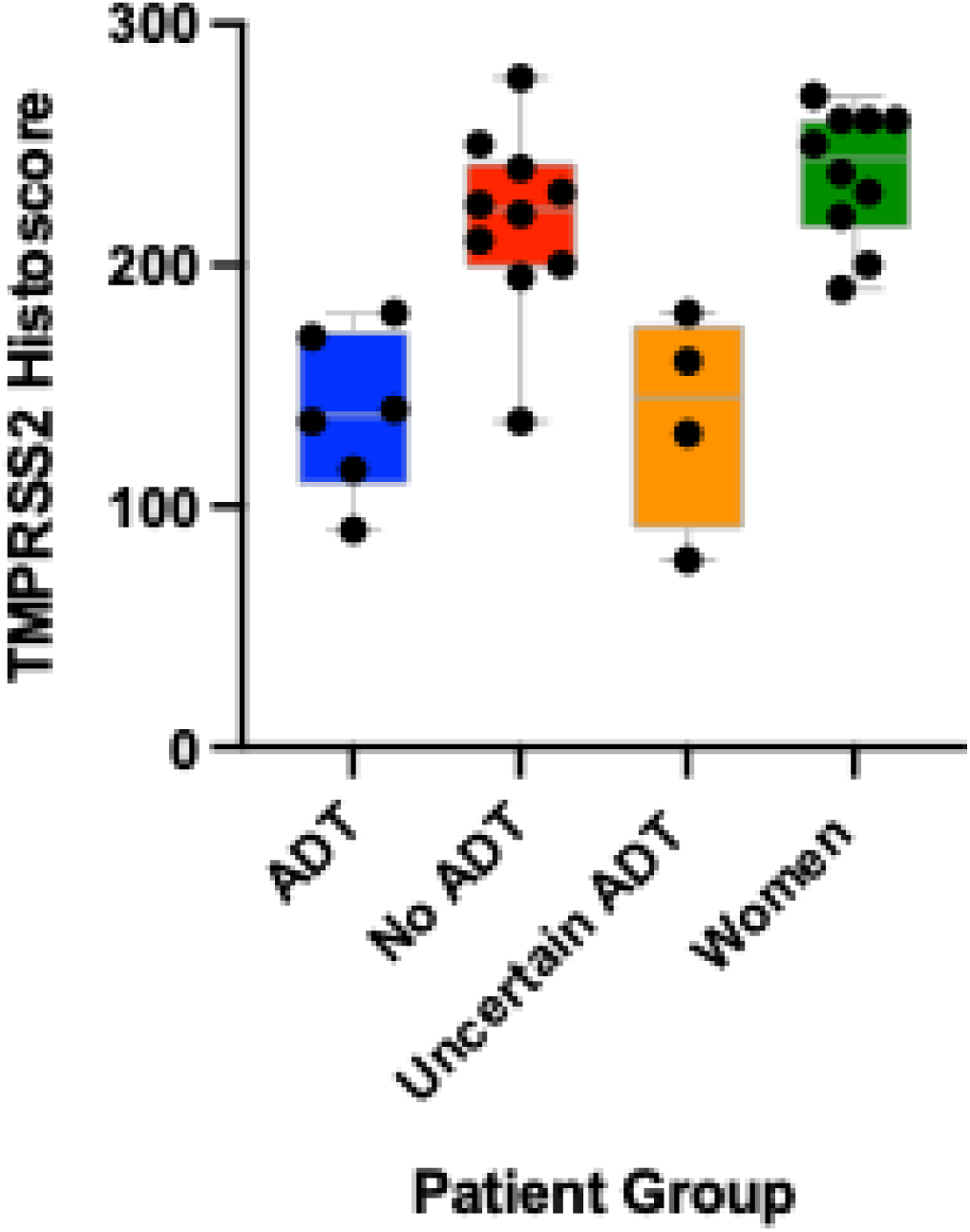
TMPRSS2 expression by patient group. Box plot showing distribution of TMPRSS2 histoscores across ADT-treated prostate cancer patients (n=6), non-ADT prostate cancer patients (n=10), patients with uncertain ADT status (n=4), and women controls (n=10). Boxes represent interquartile range, horizontal lines indicate median values, squares show means, and whiskers extend to minimum and maximum values within 1.5 × IQR. Individual data points are plotted as circles TMPRSS2 expression was significantly reduced in ADT-treated patients compared to non-ADT patients (P=.037) and women (P=.024). **Abbreviations:** ADT, androgen deprivation therapy; AR, androgen receptor; TMPRSS2, transmembrane protease serine 2.

We observed no significant correlation between TMPRSS2 expression and prostate tumor Gleason score (Figure 4). Gleason 6 cases (n=8) showed wide variation in TMPRSS2 histoscores (range: 107.5-277.5, mean: 187.5), while higher-grade tumors did not consistently exhibit higher TMPRSS2 expression. The absence of a linear relationship between Gleason grade and TMPRSS2 histoscores suggests that tumor differentiation alone does not predict pulmonary TMPRSS2 expression.

**Figure 4.**
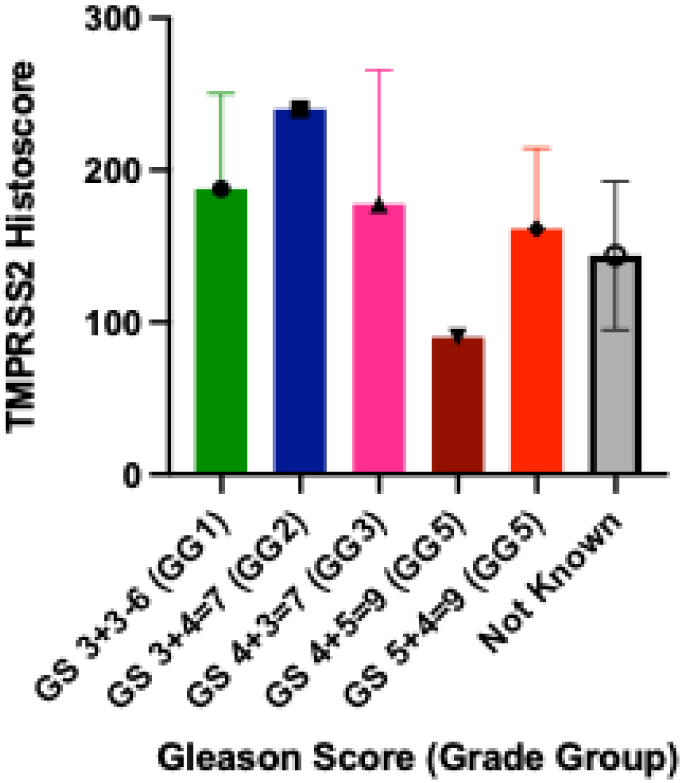
TMPRSS2 expression in lung tissue stratified by prostate tumor Gleason score. No significant correlation was observed, with Gleason 6 cases (n=8) showing wide variability (mean: 187.5; range: 107.5–277.5). Higher-grade tumors did not consistently show elevated expression, indicating that Gleason score does not predict pulmonary TMPRSS2 levels.

Similarly, no clear correlation emerged between PSA levels and TMPRSS2 expression in ADT-treated patients, though the sample size for this analysis was limited (n=5). Patients with remarkably high PSA levels (>200 ng/mL) generally exhibited lower TMPRSS2 expression (histoscores ∼90-160), while one patient with lower PSA (13.77 ng/mL) demonstrated higher expression (histoscore 240).

The presence of underlying lung disease was associated with a modest, non-significant reduction in TMPRSS2 expression (mean histoscores: 180.1 vs. 194.1, p=0.62).

### Effect of ADT Agent Type on TMPRSS2 Expression

Analysis of TMPRSS2 expression by ADT regimen revealed differential effects based on therapeutic mechanism. Direct AR antagonists (apalutamide, bicalutamide) produced the most pronounced reduction in TMPRSS2 expression, particularly when combined with GnRH modulators. Mean histoscores were lowest for bicalutamide (90) and apalutamide (115), while GnRH modulators alone or androgen biosynthesis inhibitors showed more modest reductions (mean histoscores: leuprolide 180, degarelix 170, abiraterone 135, orteronel 140) (Figure 5).

**Figure 5.**
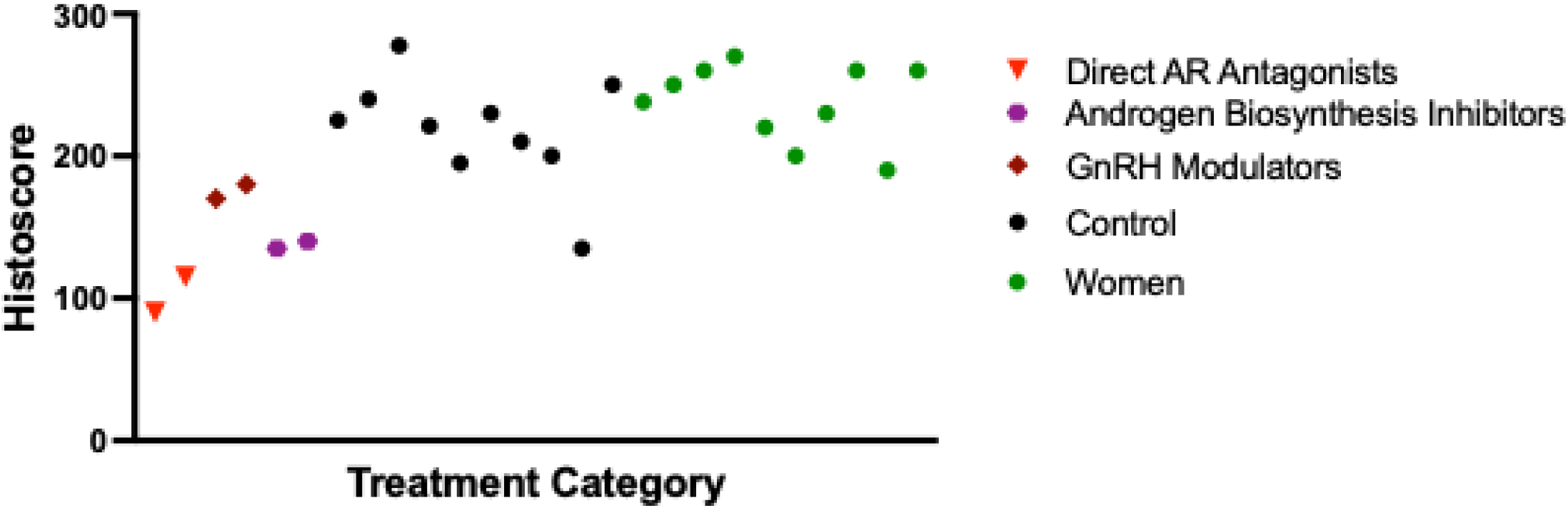
TMPRSS2 expression by specific ADT regimen. Individual patient histoscores grouped by ADT mechanism: Direct AR antagonists (bicalutamide+leuprolide, apalutamide+leuprolide), GnRH modulators (leuprolide monotherapy, degarelix), androgen biosynthesis inhibitors ( abiraterone+leuprolide, orteronel+degarelix), and control groups (no ADT, women). Direct AR antagonists produced the most pronounced reduction in TMPRSS2 expression. **Abbreviations:** ADT, androgen deprivation therapy; AR, androgen receptor; TMPRSS2, transmembrane protease serine 2. GnRH, Gonadotropin Releasing Hormone.

This pattern suggests that direct AR blockade more effectively suppresses TMPRSS2 expression than interventions targeting the hypothalamic-pituitary axis or androgen biosynthesis. The particularly marked reduction with combination therapy (direct AR antagonist plus GnRH modulator) suggests potential synergistic effects through multi-level androgen signaling suppression (Figure 6).

**Figure 6.**
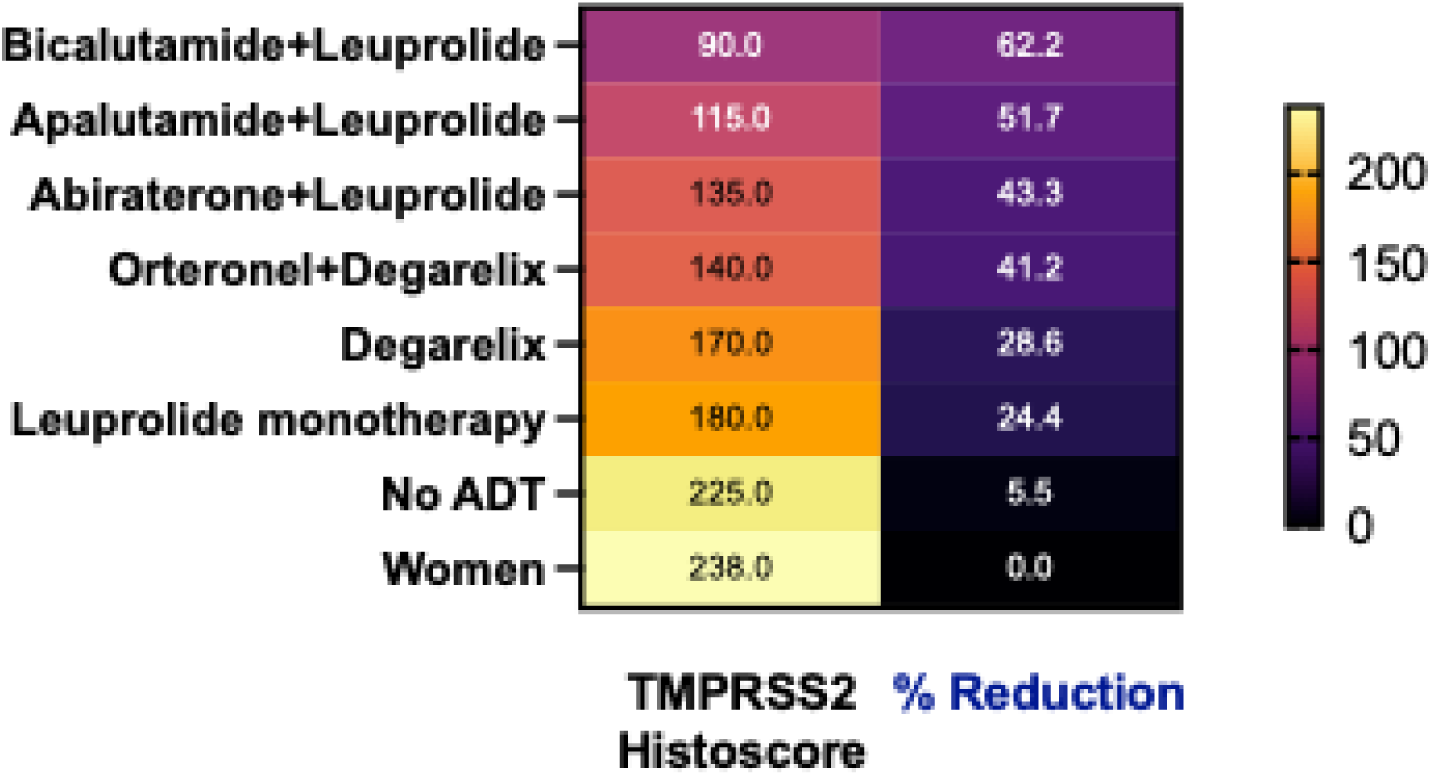
Relationship between TMPRSS2 expression and percent reduction by ADT regimen. Heat map simultaneously visualizing both absolute histoscores (left) and percent reduction from control levels (right) across ADT regimens. Darker intensity represents lower values. Direct AR antagonists (bicalutamide+leuprolide, apalutamide+leuprolide) show the lowest histoscores and highest percent reductions. **Abbreviations:** ADT, androgen deprivation therapy; AR, androgen receptor; TMPRSS2, transmembrane protease serine 2. GnRH, Gonadotropin Releasing Hormone.

## Discussion

The COVID-19 pandemic has stimulated intensive research into SARS-CoV-2 cellular entry mechanisms and potential therapeutic targets.^1,3,20,21^ TMPRSS2, a serine protease critical for S protein processing, has emerged as a key facilitator of viral entry into host cells. Upon ACE2 binding, SARS-CoV-2 undergoes conformational changes enabling TMPRSS2-mediated cleavage of the S2’ subunit, exposing the fusion peptide and initiating membrane fusion.^21^ The androgen-dependent regulation of TMPRSS2 and the male predominance in COVID-19 mortality have prompted investigations into androgen-targeted interventions as potential therapeutic strategies.

Our study provides novel evidence that androgen deprivation therapy in prostate cancer patients significantly reduces TMPRSS2 expression in lung tissue. This reduction was most pronounced with direct AR antagonists, suggesting differential efficacy based on therapeutic mechanism. These findings have several important implications for understanding COVID-19 pathogenesis and developing targeted interventions.

### Impact of ADT on TMPRSS2 Expression and COVID-19 Outcomes

Earlier studies investigating the relationship between ADT and COVID-19 outcomes have yielded mixed results. Montopoli et al. reported significantly lower SARS-CoV-2 infection rates in ADT-treated prostate cancer patients compared to untreated patients in a large Italian cohort, suggesting potential protective effects.^22^ Conversely, Shah et al. found no significant differences in hospitalization rates, oxygen requirements, or mortality between ADT-treated and untreated COVID-19 patients,^23^ while Duarte et al. observed no association between ADT use and reduced mortality in hospitalized Brazilian patients.^24^

Our findings provide a potential mechanistic explanation for these discrepancies by proving that the effect on TMPRSS2 expression varies substantially based on specific ADT regimen. Direct AR antagonists (apalutamide, bicalutamide) produced the most pronounced suppression, while GnRH modulators and androgen biosynthesis inhibitors showed more modest effects. This differential impact suggests that the protective benefit of ADT may depend on therapeutic approach, possibly explaining the heterogeneous clinical outcomes observed across studies with varied treatment protocols.

Furthermore, our observation that combination therapy (AR antagonist plus GnRH modulator) produced particularly marked TMPRSS2 suppression suggests that multi-level androgen signaling inhibition may offer enhanced protective effects. This finding aligns with recent preclinical work by Deng et al., who demonstrated that androgen receptor inhibition attenuates spike-mediated viral entry in lung and prostate cells,^25^ and Leach et al., who showed that enzalutamide decreases TMPRSS2 expression and inhibits viral entry in human lung cells and mouse models.^26^

### Therapeutic Implications and Future Directions

The significant reduction in pulmonary TMPRSS2 expression with AR-targeted therapy suggests several potential therapeutic applications. First, for prostate cancer patients at high risk of COVID-19 exposure or complications, regimens incorporating direct AR antagonists might offer dual benefits of oncologic control and potential viral protection. Second, our findings provide rationale for investigating AR antagonists as adjunctive therapy in high-risk non-cancer patients with COVID-19, particularly males with elevated androgen levels.

The differential efficacy of various ADT approaches also has important implications for clinical trial design. Future studies should stratify patients by specific ADT regimen rather than treating ADT as a homogeneous intervention. Additionally, the apparent synergistic effect of combined AR antagonism and GnRH modulation suggests that multi-target approaches may be more effective than single-agent interventions.

Our observation that direct AR blockade more effectively reduces TMPRSS2 expression than other approaches also provide insights into the molecular regulation of this key viral entry factor. This suggests that local, tissue-level androgen signaling may be more important than systemic androgen levels in regulating pulmonary TMPRSS2 expression, a finding that could inform the development of tissue-selective AR modulators with optimized pulmonary activity.

### Study Limitations and Strengths

Several limitations should be acknowledged. First, our sample size was relatively small, particularly for subgroup analyses by specific ADT regimen. Second, the retrospective nature precludes establishment of causal relationships between ADT and TMPRSS2 expression. Third, as an autopsy study, our findings may not fully reflect TMPRSS2 expression patterns in living patients with less advanced disease.

Despite these limitations, our study has important strengths. The use of matched controls and comprehensive clinicopathologic characterization allowed robust comparative analyses. The inclusion of patients on various ADT regimens permitted assessment of differential effects based on therapeutic mechanism. Finally, the direct examination of lung tissue through validated immunohistochemical methods provides more definitive evidence of TMPRSS2 expression patterns than peripheral blood markers or in vitro models.

## Conclusion

This study demonstrates that androgen deprivation therapy significantly reduces TMPRSS2 expression in lung tissue of prostate cancer patients, with direct AR antagonists producing the most pronounced effect. These findings provide a mechanistic basis for the potential protective effect of ADT against COVID-19 severity and suggest that targeted AR inhibition may represent a promising therapeutic strategy for mitigating SARS-CoV-2 infection. Future prospective studies with larger cohorts are warranted to validate these findings and assess whether TMPRSS2 suppression translates to improved clinical outcomes in COVID-19 patients.

## Data Availability

All data produced in the present work are contained in the manuscript

## Acknowledgements

We would like to acknowledge the support of the Prostate Cancer Foundation, the Pacific Northwest Prostate Cancer NIH SPORE (CA097186), Prostate Cancer Clinical Trials Consortium (PCCTC) and the U.S. Department of Defense (DOD) Prostate Cancer Research Program (PCRP), Department of Pathology & Laboratory Medicine, Oregon Health & Science University; and the Histopathology Shared Resource for pathology support (P30 CA069533 and P30 CA069533 13S5 through the OHSU-Knight Cancer Institute).

